# Alzheimer’s disease neuropathologic change mediates the relationship between ambient air pollution and dementia severity

**DOI:** 10.1101/2024.07.18.24310646

**Authors:** Boram Kim, Kaitlin Blam, Holly Elser, Sharon X. Xie, Vivianna M. Van Deerlin, Trevor M. Penning, Daniel Weintraub, David J. Irwin, Lauren M. Massimo, Corey T. McMillan, Dawn Mechanic-Hamilton, David A. Wolk, Edward B. Lee

## Abstract

**Background:** Exposure to fine particulate matter air pollution (PM_2.5_ increases risk for dementia. However, it is unknown whether this relationship is mediated by dementia-related neuropathologic change found at autopsy. We aimed to examine relationships between PM_2.5_ exposure, dementia severity, and dementia-associated neuropathologic change.

**Methods:** This cross-sectional study used harmonized demographic, clinical, genetic, and neuropathological data from autopsy cases collected from 1998 to 2022 at the Center for Neurodegenerative Disease Research brain bank, University of Pennsylvania. Cases who had common neuropathologic forms of dementia and complete data on neuropathologic measures, *APOE* genotype, and residential address were included in this study cohort. Dementia severity was measured by Clinical Dementia Rating-Sum of Boxes (CDR-SB) scores. Ten dementia-associated neuropathologic measures representing Alzheimer’s disease, Lewy body disease, limbic-predominant age related TDP-encephalopathy, and cerebrovascular disease were graded or staged according to the consensus criteria. One-year average PM_2.5_ exposure prior to death was estimated using a spatiotemporal prediction model based on residential addresses as the primary exposure measure. Linear, logistic and structural equation models were used to examine the relationships between PM_2.5_, CDR-SB and neuropathologic measures.

**Results:** A total of 861 autopsy cases were included (mean age at death 76.6 years [SD 10.3]; 481 [56%] male). Each 1 μg/m^3^ increase in one-year average PM_2.5_ concentration prior to death was associated with significantly greater cognitive and functional impairment (increase in CDR-SB score of 0.78; 95% confidence interval [CI], 0.52-1.05), faster cognitive and functional decline (change in CDR-SB scores of 0.13; 95% CI, 0.09-0.16), more severe Alzheimer’s disease neuropathologic change (ADNC; odds ratio [OR] of 1.07; 95% CI, 1.01-1.13), and a higher prevalence of large infarcts (OR, 1.17; 95% CI, 1.05-1.30). The relationship between PM_2.5_ exposure and CDR-SB was mediated by ADNC (change in CDR-SB score due to ADNC level of 0.36; 95% CI, 0.13-0.65).

**Conclusions:** PM_2.5_ exposure may increase dementia risk by increasing ADNC. Measures that improve air quality may represent a population-level intervention for the prevention of dementia.

## Introduction

Air pollution is a risk factor for dementia.^1^ Studies consistently show that exposure to air pollution, or more specifically fine particulate matter with aerodynamic diameter less than 2.5μm (PM_2.5_), is associated with an increased incidence of dementia,^2^ impaired cognitive function,^3,4^ and accelerated cognitive decline.^5^ Disease-modifying therapies that target the underlying neuropathology of Alzheimer’s disease (AD) show partial clinical benefit, but substantial economic and logistical barriers preclude widespread implementation and serious side effects occur in some instances.^6,7^ Efforts to curb PM_2.5_ levels has the potential to benefit health outcomes and prevent dementia risk at the population level.

Despite growing evidence of the adverse effect of PM_2.5_ on cognition,^8,9^ the underlying biological mechanisms for this relationship are largely unknown. PM_2.5_ exposure has been associated with brain volume loss,^10^ cortical atrophy,^11,12^ lower cerebrospinal fluid amyloid-β_42_ levels^13^, increased amyloid PET positivity,^14^ and accelerated epigenic aging,^15^ raising the possibility that PM_2.5_ might contribute to dementia risk by enhancing Alzheimer’s disease neurodegenerative disease pathways.^16,17^ Post-mortem autopsy examination remains the gold standard for the neuropathologic diagnosis of Alzheimer’s disease and related dementias. However, few post-mortem studies have investigated the association between PM_2.5_ and neurodegenerative disease pathology^18,19^, and the tripartite relationship between PM_2.5_, neurodegenerative disease pathology, and cognitive and functional outcomes has not been studied.

To address this knowledge gap, we studied here the relationships between PM_2.5_ exposure, neuropathologic change, and cognitive and functional impairment in a large, well-characterized autopsy cohort. Using high spatial resolution estimates of PM_2.5_ obtained from a validated prediction model,^20^ we determined whether PM_2.5_ exposure was associated with dementia severity measured by CDR-SB scores to assess cognitive and functional performance in a well-characterized autopsy cohort. We further evaluated whether PM_2.5_ exposure was associated with differences in the burden of the most common dementia-related neuropathologies including Alzheimer’s disease neuropathologic change (ADNC), Lewy body disease (LBD), limbic-predominant age-related TDP-43 encephalopathy neuropathologic change (LATE-NC), and cerebrovascular disease.^21^ Finally, we evaluated whether the effect of PM_2.5_ on dementia severity was mediated by neuropathologic change.

## Methods

### Study design and participants

We obtained harmonized demographic, clinical, and neuropathology data for autopsy cases from the Integrated Neurodegenerative Disease (INDD) database at the Center for Neurodegeneration Disease Research (CNDR) at the University of Pennsylvania as previously described.^22^ Cases were recruited mainly through the Penn Alzheimer’s Disease Research Center which focuses on dementia of the Alzheimer’s type (as opposed to vascular or other dementias) and the Penn Parkinson’s Disease and Movement Disorder Center which includes a research cohort focusing on cognitive dysfunction in the setting of LBD. Details on the study cohort are provided in the appendix p 3 and 5. In brief, we restricted our autopsy cases to individuals older than the age of 40 with common neuropathological forms of dementia from 1998 through 2022. We excluded cases with missing neuropathology, *APOE* ε4 allele status, and/or residential address data resulting in a cohort of 861 cases.

### Demographic and clinical assessment

Self-reported sex, race, ethnicity, and years of education, and physician-reported brain weight and *APOE* ε4 allele status were extracted from the INDD database. Dementia severity was based on Clinical Dementia Rating-Sum of Boxes (CDR-SB) scores, used as a continuous variable. Age at the last CDR-SB testing and the number of years between the last CDR-SB test and death were also obtained.

### Neuropathological assessment

Using consensus criteria,^23–26^ ten neuropathologic summary scores were obtained, representing four proteinopathies (tau, β-amyloid, α-synuclein, and TDP-43) and three cerebrovascular lesions (infarcts, amyloid angiopathy, arteriolosclerosis) for each case as follows: Thal amyloid phase (A scores from 0 to 3 representing Thal amyloid phase 0, 1/2, 3, and 4/5), Braak stage (B scores from 0 to 3 representing Braak stage 0, I/II, III/IV, and V/VI), CERAD score (C scores from 0 to 3 representing absent, mild, moderate, and severe neocortical neuritic plaque burden), level of ADNC (from 0 to 3 representing not, low, intermediate, and high level),^24^ absence/presence of LBD (0, no Lewy pathology or brainstem LBD or 1, amygdala-predominant, limbic/transitional, or neocortical/diffuse LBD),^23^ LATE-NC stage (from 0 to 3),^25^ absence/presence of large infarcts, absence/presence of moderate to high occipital cerebral amyloid angiopathy burden, absence/presence of moderate to high arteriolosclerosis in occipital white matter, and likelihood that cerebrovascular pathology contributed to cognitive impairment (VCING scores from 1 to 3 representing low, intermediate, and high likelihood).^26^ Further details are described in appendix p 4..

### Air pollution exposure assessment

Ambient PM_2.5_ exposure was estimated using annual average PM_2.5_ concentrations across the US at a 0.01°×0.01° (approximately 1.1km x 1.1km) grid cell resolution from 1998 to 2022, by combining satellite retrievals, chemical transport modelling and ground-based observations (V5.GL.04). These data were recently updated based on previously described data sets^20^ and are publicly available through the Washington University, St. Louis. We then spatially matched each case’s geocoded residential addresses before death to grid cells to obtain one-year average PM_2.5_ exposures prior to death as a continuous variable (appendix p 4).

### Statistical analysis

We used linear regression to estimate the cross-sectional association of one-year average PM_2.5_ concentrations prior to death with CDR-SB scores at last assessment prior to death, adjusting for sex, age at CDR-SB assessment, race, and years of education. For the subset of cases with multiple CDR-SB records, we used a linear mixed-effects model to estimate an adjusted association of PM_2.5_ concentrations with longitudinal change in CDR-SB scores.

Ordinal logistic regression models were used to estimate the association between PM_2.5_ concentration and ordered categorical outcomes that included Thal amyloid phase, Braak stage, CERAD score, ADNC level, LATE-NC stage, and VCING likelihood level. Binary logistic regression models were used to estimate the association between PM_2.5_ concentration and the presence/absence of LBD, large infarcts, occipital lobe cerebral amyloid angiopathy, and occipital white matter arteriolosclerosis. These models were adjusted for sex, age at death, and *APOE* ε4 allele status.

To explore the tripartite relationship between PM_2.5_ concentration, CDR-SB scores, and neuropathologic change as an ordered categorical mediator, we performed a mediation analysis with structural equation modelling using the R “lavaan” package.^27^ PM_2.5_ exposure was the exogenous independent variable, ADNC and large infarcts were potential endogenous mediators, CDR-SB was the endogenous dependent variable, and sex, age at CDR assessment, interval years between CDR assessment and death, race, years of education, and *APOE* ε4 allele status were exogenous control variables. In this model, age at death was not included as a control variable as it showed a perfect multicollinearity with a linear combination of age at CDR assessment and interval years between CDR assessment and death variables. We estimated indirect, direct, and total effects using a bias-corrected, accelerated bootstrap confidence interval method with 2000 simulations. Dichotomous nuisance variables were encoded as follows: sex (0, female, reference group; 1, male), race (0, non-white, reference group; 1, white), *APOE* ε4 allele status (0, no *APOE* ε4 (-), reference group; 1, *APOE* ε4 (+)). Diagnostic tests were performed to assess model fitness (appendix p 4).

To explore the robustness of our findings, additional sensitivity analyses were performed using additional exposure time windows corresponding to 3-year and 5-year average PM_2.5_ exposure before their death. Analyses were performed with R version 4.2.3. All statistical tests were two-sided.

## Results

### Cohort characteristics

Demographic, clinical, and neuropathological features of the cohort are provided in table 1. Within the cohort, 481 (55.9%) were male, the mean (SD) age of death was 76.6 (10.3) years, most of the subjects were white (92.4%) and non-Hispanic or Latino (98.7%), and 49.4% carried at least one ε4 allele of the *APOE* which is the strongest genetic risk factor for sporadic AD. Overall, our cohort was highly educated with an average of 15.3 years of education, with a moderate level of dementia corresponding to an average CDR-SB score of 10.0 at last assessment. Annual average change in CDR-SB scores was 1.6 with an average assessment interval of 5 years. The most common clinical diagnoses were AD (36.7%) and Parkinson’s disease with dementia (15.7%).

**Table 1.** Characteristics of study participants.

| Characteristic | No. of Subjects | % or mean±SD |
| --- | --- | --- |
| **Demographic** |  |  |
| Sex | 861 |  |
| Male | 481 | 55.9 |
| Female | 380 | 44.1 |
| Age at death - yr | 861 | 76.6±10.3 |
| Race <sup>a</sup> | 847 |  |
| White | 783 | 92.4 |
| Non-White | 64 | 7.6 |
| Ethnicity <sup>a</sup> | 764 |  |
| Not Hispanic or Latino | 754 | 98.7 |
| Hispanic or Latino | 10 | 1.3 |
| Brain weight - g | 858 | 1211.6±171.8 |
| *APOE* ε4 allele status | 861 | |
| *APOE* ε4 (+) | 425 | 49.4 |
| ε2/ε4 | 20 | 2.3 |
| ε3/ε4 | 307 | 35.7 |
| ε4/ε4 | 98 | 11.4 |
| *APOE* ε4 (-) | 436 | 50.6 |
| ε2/ε2 | 3 | 0.3 |
| ε2/ε3 | 64 | 7.4 |
| ε3/ε3 | 369 | 42.9 |
| Education - yr | 616 | 15.3±3.1 |
| **Clinical** |  |  |
| CDR-SB at last assessment <sup>b</sup> | 348 | 10±6.4 |
| Age at last CDR-SB assessment -yr | 348 | 76.1±10.7 |
| Interval between last CDR-SB assessment and death -yr | 348 | 2.9±3 |
| Annual change in CDR-SB | 263 | 1.6±1.9 |
| Primary clinical diagnosis |  |  |
| Possible/Probable Alzheimer's disease | 316 | 36.7 |
| Parkinson's disease with dementia | 135 | 15.7 |
| Neurologically normal | 111 | 12.9 |
| Parkinson's disease | 72 | 8.4 |
| Dementia with Lewy bodies | 67 | 7.8 |
| Frontotemporal degeneration | 48 | 5.6 |
| Corticobasal syndrome | 33 | 3.8 |
| Logopenic primary progressive aphasia | 16 | 1.9 |
| Mild cognitive impairment | 13 | 1.5 |
| Vascular dementia | 12 | 1.4 |
| Dementia, uncertain etiology | 9 | 1 |
| Posterior cortical atrophy | 7 | 0.8 |
| Other | 22 | 2.6 |
| **Neuropathological <sup>c</sup>** |  |  |
| Thal amyloid phase | 861 |  |
| 0 | 82 | 9.5 |
| 1/2 | 126 | 14.6 |
| 3 | 129 | 15.0 |
| 4/5 | 524 | 60.9 |
| Braak NFT stage | 861 |  |
| 0 | 34 | 3.9 |
| I/II | 225 | 26.1 |
| III/IV | 150 | 17.4 |
| V/VI | 452 | 52.5 |
| CERAD score | 861 |  |
| 0 | 197 | 22.9 |
| 1 | 69 | 8.0 |
| 2 | 122 | 14.2 |
| 3 | 473 | 54.9 |
| ADNC level | 861 |  |
| Not | 81 | 9.4 |
| Low | 214 | 24.9 |
| Intermediate | 130 | 15.1 |
| High | 436 | 50.6 |
| Lewy pathology | 861 |  |
| No or brainstem predominant | 404 | 46.9 |
| Amygdala, transitional/limbic,<br>of diffuse/neocortical predominant | 457 | 53.1 |
| LATE | 861 |  |
| No | 602 | 69.9 |
| LATE-NC stage 1 | 54 | 6.3 |
| LATE-NC stage 2 | 176 | 20.4 |
| LATE-NC stage 3 | 29 | 3.4 |
| Large infarct | 861 |  |
| No | 813 | 94.4 |
| Yes | 48 | 5.6 |
| Cerebral amyloid angiopathy<br>burden in occipital lobe | 861 |  |
| Absent to mild | 626 | 72.7 |
| Moderate to severe | 235 | 27.3 |
| Arteriolo sclerosis burden in<br>occipital white matter | 861 |  |
| Absent to mild | 717 | 83.3 |
| Moderate to severe | 144 | 16.7 |
| VCING level | 861 |  |
| Low | 770 | 89.4 |
| Intermediate | 68 | 7.9 |
| High | 23 | 2.7 |

Abbreviations: NFT, neurofibrillary tangle; CERAD, Consortium to Establish a Registry for Alzheimer's Disease; ADNC, Alzheimer's disease neuropathologic change; LATE, limbic-predominant age-related TDP-43 encephalopathy; VCING, vascular cognitive impairment neuropathology guidelines.
<sup>a</sup> Race and ethnicity were reported by the study subjects.
<sup>b</sup> Scores on the Clinical Dementia Rating–Sum of Boxes (CDR-SB) at last assessment prior to death range from 0 to 18, with higher scores indicating greater cognitive and functional impairment.
<sup>c</sup> All pathological measures were recorded according to consensus criteria.

More than a half of the cohort exhibited severe AD neuropathologic measures including Thal amyloid phase 4/5 (60.9%), Braak stage V/VI (52.5%), CERAD score 3 (54.9%), and high overall ADNC (50.6%). 53.1% exhibited LBD, and 30.1% exhibited LATE-NC. Notably, cerebrovascular disease was not a common feature of this autopsy cohort with relatively few cases exhibiting large infarcts (5.6%), moderate to severe occipital lobe cerebral amyloid angiopathy (27.3%), moderate to severe occipital white matter arteriolosclerosis burden (16.7%), or overall high likelihood that cerebrovascular pathology contributed to cognitive impairment (2.7%).

### Air pollution and cognition

To determine whether this autopsy cohort was reasonably similar to epidemiologic observations that link PM_2.5_ exposure to impaired cognitive and functional performance, we spatiotemporally analyzed PM_2.5_ data for the study cohort based on geocoded addresses and the year of death ranging from 1998 to 2022 (appendix p 6 – 8). One-year average PM_2.5_ exposures before death was 10.1 μg/m^3^, ranging from 5 to 18.6 μg/m^3^. The average PM_2.5_ concentration decreased by 51.8% from 2000 through 2022, which was highly correlated with the 42.2% decrease in PM_2.5_ concentrations across the entire US over the same period (n=23, r = 0.97, and p < 0.001 by Spearman’s rank correlation; appendix p 7 - 8).

Each 1 μg/m^3^ increase in one-year average exposure to PM_2.5_ before death was associated with a 0.78 point increase in the last CDR-SB score before death (95% confidence interval [CI], 0.52-1.05), indicating that more air pollution was associated with worse cognitive and functional impairment. This relationship appeared to be independent of the adjusted protective effect of education on cognitive and functional impairment, where each one-year increment in educational attainment was associated with a 0.31 point decrease in last CDR-SB score prior to death (95% CI, -0.53 - -0.09; table 2).

**Table 2.** PM_2.5_ exposure and Clinical Dementia Rating–Sum of Boxes (CDR-SB).^a^.

| **Exposure** | **Estimate (95% CI)<sup>b</sup>** | **P Value<sup>c</sup>** |
| --- | --- | --- |
| No. of participants (N)=348<sup>d</sup> |  |  |
| PM<sub>2.5</sub> | 0.78 (0.52 - 1.05) | <0.001 |
| Education | -0.31 (-0.53 - -0.09) | 0.005 |

Abbreviation: CI, confidence interval.
<sup>a</sup> Associations were evaluated by a linear regression model in which the outcome variable was Clinical Dementia Rating-Sum of Boxes (CDR-SB) score at last assessment with higher values indicating greater cognitive and functional impairment, the exposure variables were one-year average PM<sub>2.5</sub> exposure before death and years of education, and covariates were sex, age at last CDR-SB assessment, and race.
<sup>b</sup> Estimated effects and 95% confidence intervals were shown adjusted changes in scores on CDR-SB at last assessment prior to death per 1 µg/m<sup>3</sup> increase in one-year average PM<sub>2.5</sub> exposure before death and one-year increase in educational attainment, with positive values indicating greater cognitive and functional impairment.
<sup>c</sup> P values less than 0.05 were considered statistically significant.
<sup>d</sup> 348 cases with CDR-SB data were included in this analysis.

In addition, each 1 μg/m^3^ increase in one-year average exposure to PM_2.5_ before death was associated with a 0.13 point increase in annual change in CDR-SB scores (95% CI, 0.09-0.16; table 3), indicating that more air pollution was associated with faster cognitive and functional decline.

**Table 3.** PM_2.5_ exposure and longitudinal change in Clinical Dementia Rating–Sum of Boxes (CDR-SB). ^a^.

| **Exposure** | **Estimate (95% CI)<sup>b</sup>** | **P Value<sup>c</sup>** |
| --- | --- | --- |
| No. of participants=236<sup>d</sup> |  |  |
| Interaction term between PM<sub>2.5</sub> and interval years between CDR-SB assessment and death | 0.13 (0.09, 0.16) | <0.001 |

Abbreviation: CI, confidence interval.
<sup>a</sup> The association was evaluated by a linear mixed-effects model in which the outcome variable was annual change in Clinical Dementia Rating–Sum of Boxes (CDR-SB) scores obtained from multiple assessments with higher values indicating faster cognitive and functional impairment, the exposure variable was the interaction term between one-year average PM<sub>2.5</sub> level exposed to study participants before their death and interval years between last CDR-SB assessment and death, and fixed covariates were interval years between last CDR-SB assessment and death, sex, age at last CDR-SB assessment, race, and years of education.
<sup>b</sup> The estimated effect and 95% confidence interval were shown an adjusted annual change in scores on CDR-SB at last assessment prior to death per 1 µg/m<sup>3</sup> increase in one-year average PM<sub>2.5</sub> exposure before death, with positive values indicating faster cognitive and functional impairment.
<sup>c</sup> P value less than 0.05 was considered statistically significant.
<sup>d</sup> 236 cases with multiple CDR-SB data were included in this analysis.

### Air pollution and neuropathologic change

The adjusted relationship between PM2.5 exposure and dementia-related neuropathologies was examined. Every 1 μg/m^3^ increase in one-year average exposure to PM_2.5_ before death was associated with overall an increased odds ratio for worse neuropathologic outcomes including higher Thal amyloid phase (odds ratio [OR], 1.07; 95% CI, 1.01-1.13), Braak stage (OR, 1.07; 95% CI, 1.02-1.13), CERAD score (OR, 1.11; 95% CI, 1.05-1.17), level of ADNC (OR, 1.06; 95% CI, 1.01-1.12), and prevalence of large infarcts (OR, 1.17; 95% CI, 1.05-1.30). In contrast, PM_2.5_ was not associated with LBD stage (OR, 1.01; 95% CI, 0.96-1.06), LATE-NC stage (OR, 1.02; 95% CI, 0.96-1.08), prevalence of moderate or severe occipital lobe cerebral amyloid angiopathy burden (OR, 0.98; 95% CI, 0.92-1.04), occipital white matter arteriolosclerosis burden (OR, 0.99; 95% CI, 0.92-1.06), and overall VCING level (OR, 1.05; 95% CI, 0.97-1.14) (figure. 1).

**Figure 1.**
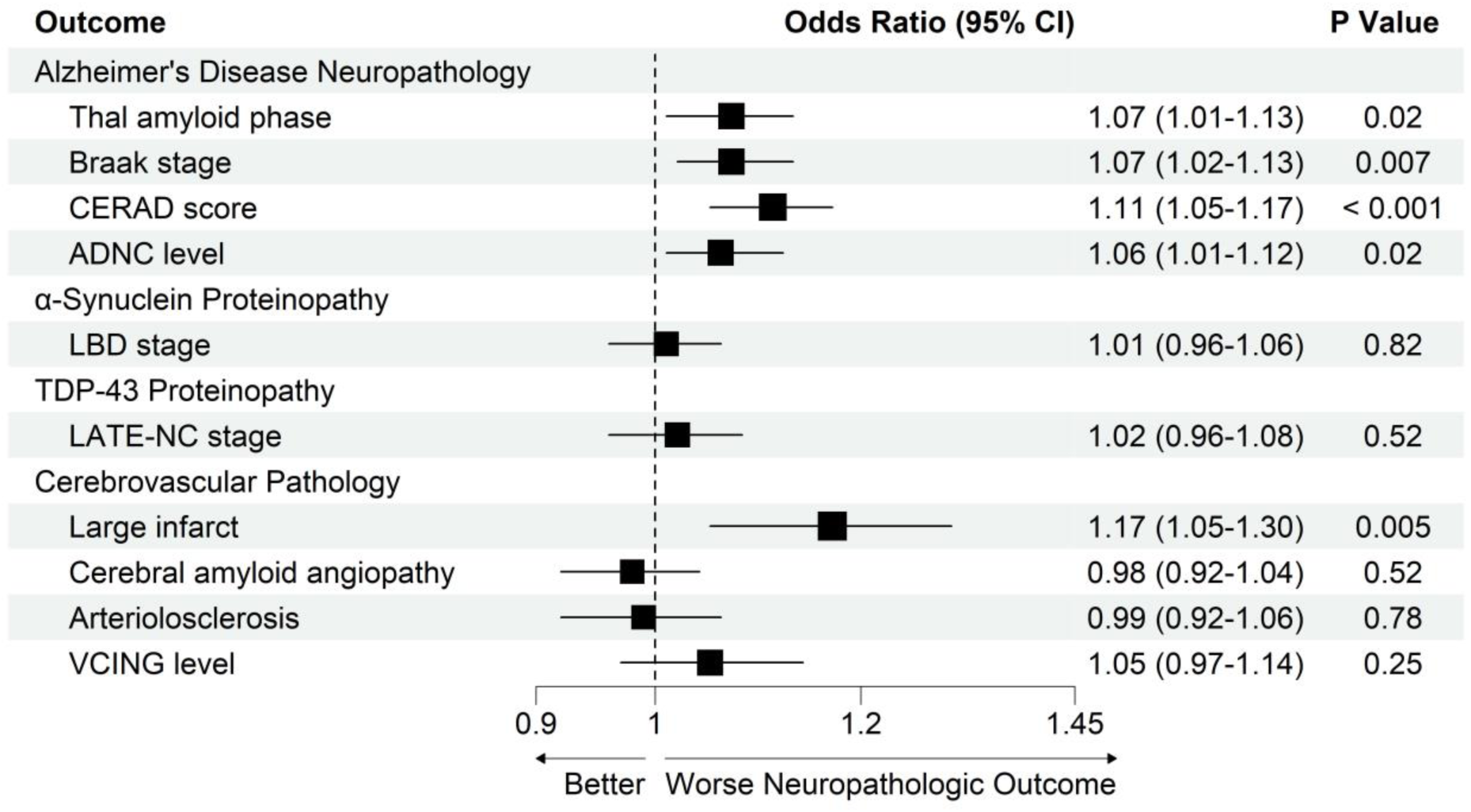
PM_2.5_ exposure and dementia-related neuropathologies. Abbreviations: CERAD, Consortium to Establish a Registry for Alzheimer’s Disease; ADNC, Alzheimer’s disease neuropathologic change; LBD, Lewy body disease; LATE, limbic-predominant age-related TDP-43 encephalopathy; VCING, vascular cognitive impairment neuropathology guidelines; CI, confidence interval. Forest plot of odds ratios and 95% confidence intervals for neuropathologic outcomes corresponding to every 1 μg/m^3^ increase in one-year average PM_2.5_ exposure before death for study cases. Odds ratios and 95% confidence intervals are reported from a series of ordinal logistic regression models with one-year average PM_2.5_ before death as the exposure variable and ordinal Thal amyloid phase, ordinal Braak stage, ordinal CERAD score, ordinal ADNC level, ordinal LATE-NC stage, and ordinal VCING score as outcome variables, and binary logistic regression models with one-year average PM_2.5_ exposure before death as the exposure variable and dichotomously treated LBD stage, presence of large infarcts, presence of occipital cerebral amyloid angiopathy, and presence of arteriolosclerosis as outcome variables. The odds ratios and 95% confidence intervals greater than 1 indicate worse neuropathologic outcomes. All models were controlled for sex, age at death, and *APOE* ε4 status. 861 cases with non-missing values were used for all analyses. P values less than 0.05 were considered statistically significant.

### Air pollution, neuropathologic change, and dementia

Based on the findings above that exposure to higher PM_2.5_ levels was associated with increased CDR-SB scores (table 2), increased ADNC, and increased cerebrovascular disease (figure 1), we examined whether the association between PM_2.5_ and CDR-SB was mediated by ADNC or the presence of large infarcts. A parallel mediation analysis with structural equation modelling revealed a significant mediation effect of ADNC (indirect effect; estimated change in CDR-SB scores due to increasing ADNC level, 0.36; 95% CI, 0.13-0.65) on the association between PM_2.5_ exposure and CDR-SB scores, where about 63% of the estimated total effect of PM_2.5_ exposure on CDR-SB scores (estimated change in CDR-SB scores, 0.57; 95% CI, 0.35-0.81) was mediated via ADNC. The presence of large infarcts was not a statistically significant mediator (indirect effect; estimated change in CDR-SB scores due to increasing prevalence of large infarcts, 0.11; 95% CI, -0.11 - 0.87), and the estimated direct effect of PM_2.5_ on CDR-SB independent from ADNC and the presence of large infarcts was not significant (estimated change in CDR-SB scores independent from ADNC level and the presence of large infarcts, 0.10; 95% CI, -0.48 - 0.72). This adjusted model yielded adequate model fit indices (comparative fit index, 0.895; root mean square error of approximation, 0.088; Tucker-Lewis index, 0.965) (figure. 2).

**Figure 2.**
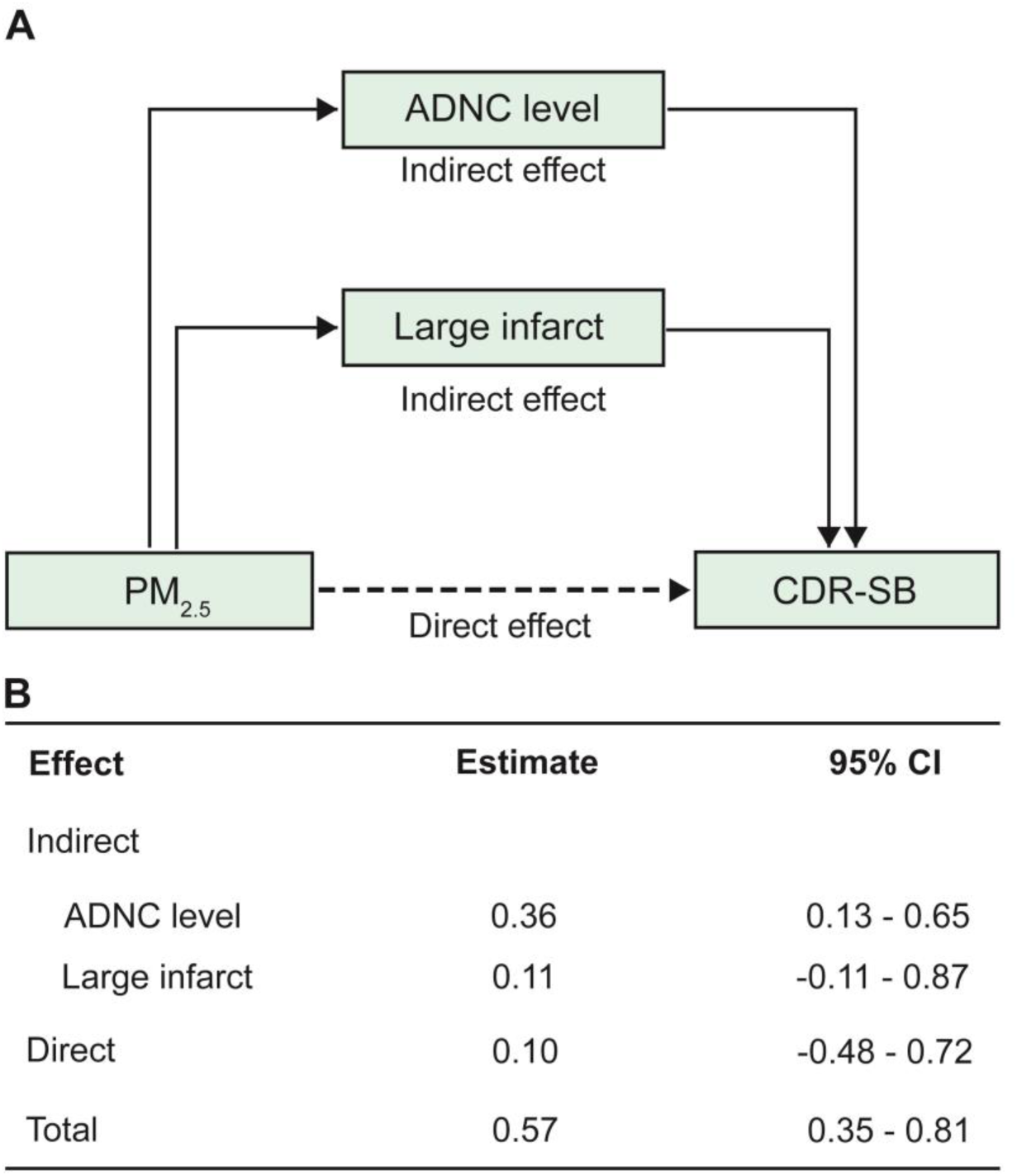
Alzheimer’s disease neuropathologic change mediates the relationship between PM_2.5_ exposure and Clinical Dementia Rating–Sum of Boxes (CDR-SB) Abbreviations: ADNC, Alzheimer’s disease neuropathologic change; CI, confidence interval. Panel A displays a path diagram of the structural equation modelling for relationships among PM_2.5_ exposure, ADNC or the presence of large infarcts, and CDR-SB with one-year average PM_2.5_ exposure before death as the exposure variable, ordinal ADNC level and presence of infarcts as parallel mediators, and CDR-SB at last assessment as the outcome variable. In this model, indirect effects are the effect of PM_2.5_ exposure on CDR-SB through ADNC (solid arrow, top) or the presence of large infarcts (solid arrow, bottom) and the direct effect is the effect of PM_2.5_ exposure on CDR-SB that is independent from mediators such as ADNC or presence of large infarcts (dotted arrow). Panel B shows estimates of indirect, direct, and total effects with 95% confidence intervals from mediation analysis based on structural equation modelling using a bias-corrected and accelerated bootstrap confidence interval method with 2000 simulations. Positive values indicate greater cognitive and functional impairment. This model was controlled for sex, age at death, interval years between last CDR-SB assessment and death, race, years of education, and *APOE* ε4 status as confounding variables. 348 cases with CDR-SB data were used for this analysis.

### Additional Sensitivity Analyses

Post hoc sensitivity analyses using expanded PM_2.5_ exposure time frames of either 3- or 5-years prior to death were largely similar to the results obtained from our primary analysis with the exception that 5-year average PM_2.5_ exposure before death was associated with a marginally decreased odds ratio for a higher prevalence of moderate or severe burden of occipital lobe cerebral amyloid angiopathy (appendix p 9 - 13).

## Discussion

In this relatively large autopsy series, we found that exposure to higher PM_2.5_ levels was associated with significantly greater and faster cognitive and functional impairment. Higher PM_2.5_ concentrations were also associated with more severe amyloid and tau pathologies, culminating in more advanced overall ADNC. Higher PM_2.5_ exposure was also associated with a higher prevalence of large infarcts. Most importantly, the impact of PM_2.5_ on dementia severity was largely mediated by an increase in ADNC. These results suggest that PM_2.5_ may directly affect brain vulnerability where increased ADNC appears to mediate PM_2.5_-induced cognitive dysfunction. These associations were not modified by sex or *APOE* ε4 allele status.

Our findings are consistent with prior population-based cohort studies which have demonstrated cross-sectional associations between PM_2.5_ exposure and cognitive decline.^3,11^ Moreover, antemortem human studies consistently show that PM_2.5_ exposure is associated with increased AD biomarkers including structural MRI changes, cerebrospinal fluid measures,^13^ and amyloid PET outcomes.^14,28–30^ These support the hypothesis that PM_2.5_ exposure is deleterious for brain maintenance pathways, resulting in worsening of ADNC.^31,32^

Two prior studies examining the effects of PM_2.5_ on neurodegenerative disease pathology have been reported, one of which demonstrated a similar association between higher PM_2.5_ exposure and higher ADNC (CERAD score) based on study of an Alzheimer’s Disease Research Center autopsy cohort.^19^ In contrast, no significant effect of PM_2.5_ exposure and amyloid, tau or overall ADNC was observed in the second study of a community-based autopsy cohort.^18^ This inconsistency may be related to differences in autopsy cohort characteristics. Our cohort consisted mainly of symptomatic dementia cases that were enriched for AD dementia, perhaps allowing for more homogeneity and therefore statistical power to detect Alzheimer’s disease specific effects and in contrast with community-based cohorts where the causes of cognitive dysfunction are likely to be more heterogeneous. In contrast with these prior studies, we included individual cases’ cognitive and functional testing data which revealed that the relationship between PM_2.5_ exposure and cognitive and functional dysfunction was largely mediated by increased ADNC.

One strength of this study is the comprehensive study of a large autopsy series with well-defined demographical, clinical, and neuropathological profiles based on consensus criteria. Furthermore, we used high resolution prediction data to estimate individual-level PM_2.5_ exposure at each residential location, and the air pollution exposure within this cohort was similar to overall trends in PM_2.5_ concentrations across the country. Finally, we demonstrated the robustness of our findings with sensitivity analyses demonstrating that similar results were obtained when using 1-, 3- or 5-years PM_2.5_ exposure before death.

Despite these strengths, we note the following limitations. First, the vast majority of subjects in our cohort were white and/or non-Hispanic or Latino, so our findings cannot be extrapolated to general population and are limited in terms of finding potential modifying effects of race and/or ethnicity. Second, due to the cross-sectional nature of autopsy studies, we were limited to fully examine longitudinal effects of air pollution on neuropathologic outcomes. Third, our autopsy cohort included a large proportion of cases through research programs that did not enroll individuals with vascular dementia. This selection bias resulted in very few cases in this cohort with severe cerebrovascular pathology, likely underestimating associations with PM_2.5_ of cerebrovascular disease. Indeed, there is strong epidemiologic evidence linking PM_2.5_ exposure and stroke.^33,34^ Finally, in this retrospective approach, we cannot comprehensively evaluate potential confounding factors, including differences in longitudinal exposures, that may affect downstream neuropathologic change and cognition.

## Conclusions

This autopsy cohort study reinforces the finding that PM_2.5_ exposure appears to negatively affect cognitive function and suggests that this relationship may be mediated by ADNC. Our findings suggest that PM_2.5_ exposure may exacerbate AD pathogenesis. Thus, efforts to improve air quality may benefit the population by diminishing risk for Alzheimer’s disease neuropathology.

### Contributors

BK and EBL conceptualized and designed the study, did statistical analysis, and drafted the Article. All authors gathered and interpreted data, assisted with revisions, and approved the final version. EBL supervised the study.

## Data Availability

All data produced in the present study are available upon reasonable request to the authors.

## Acknowledgments

This work was funded by the US National Institutes of Health (P30AG072979, P01AG066597, U19AG062418, P30ES013508). We thank the patients and their families for contributing to our research and for participating in the brain donation program. We are indebted to John Q. Trojanowski for his contributions which persist posthumously.

