## Supplementary Appendix for "Alzheimer’s disease neuropathologic change mediates the relationship between ambient air pollution and dementia severity"

Author affiliations:
[1] Translational Neuropathology Research Laboratory, Department of Pathology and Laboratory Medicine, Perelman School of Medicine at the University of Pennsylvania, Philadelphia, PA 19104, USA

[2] Department of Neurology, Perelman School of Medicine at the University of Pennsylvania, Philadelphia, PA, 19104, USA

[3] Department of Biostatistics, Epidemiology and Informatics, Perelman School of Medicine at the University of Pennsylvania, Philadelphia, PA 19104, USA

[4] Center for Neurodegenerative Disease Research, Department of Pathology and Laboratory Medicine, Perelman School of Medicine at the University of Pennsylvania, Philadelphia, PA 19104, USA

[5] Center of Excellence in Environmental Toxicology, Department of Systems Pharmacology and Translational Therapeutics, Perelman School of Medicine at the University of Pennsylvania, Philadelphia, PA 19104, USA

[6] Department of Psychiatry, Perelman School of Medicine at the University of Pennsylvania, Philadelphia, PA, 19104, USA

[*] Corresponding Author: Edward B. Lee, MD, PhD
Director, Center for Neurodegenerative Disease Research Brain Bank
Co-Director, Institute of Aging
Associate Director, Penn Alzheimer's Disease Research Center
Associate Professor of Pathology and Laboratory Medicine
Perelman School of Medicine at the University of Pennsylvania
613A Stellar Chance Laboratories
422 Curie Blvd
Philadelphia, PA 19104, USA


Mediation Analysis of Dementia-Related Neuropathologic Change on the Association between Different $PM_{2.5}$ Exposure Time Windows and CDR-SB (Sensitivity Analysis)………..…page 13

References……………………………………………………………………………….page 14

### Study cohort

The Center for Neurodegeneration Disease Research (CNDR) brain bank has been collecting autopsy cases since 1985.[1] These are mainly from participants enrolled in neurodegenerative research clinical cores at the University of Pennsylvania and are also from patients from the University of Pennsylvania Health System. Participants from the neurodegenerative research programs gave written informed consent for autopsy antemortem, and consent for autopsy was obtained from next-of-kin at time of death.

In this autopsy-based cross-sectional study, we obtained data on demographic, clinical, and neuropathological features for autopsy cases from the Integrated Neurodegenerative Disease (INDD) database at the Center for Neurodegeneration Disease Research (CNDR), as previously described.[1] Out of 2041 autopsy cases, we included cases older than the age of 40 that obtained from 1998 from 2022. To restrict study subjects to cases with common neuropathological forms of dementia including normal aging, we excluded cases who had a primary neuropathological diagnosis of rare neurodegenerative diseases such as amyotrophic lateral sclerosis, argyrophilic grain disease, corticobasal degeneration, Creutzfeldt–Jakob disease, Down syndrome, frontotemporal lobar degeneration, globular glial tauopathy, multiple system atrophy, Pick's disease, and progressive supranuclear palsy. Next, we excluded cases with incomplete/missing neuropathological evaluation due to limited tissue availability, or missing *APOE* genotype data. After excluding cases with missing residential addresses, 861 cases were included in this study cohort.

### Neuropathological assessment

Macroscopic and microscopic findings based on hematoxylin and eosin (H&E) staining and immunohistochemical staining against tau (PHF-1), amyloid-β (NAB228), α-synuclein (Syn303), and TDP-43 (ID3) of sixteen brains regions.[1] Each brain region was evaluated for the severity of each pathological lesion using semi-quantitative scoring (0, absent; 0.5, rare; 1, mild; 2, moderate; 3, severe) and reviewed by board-certified neuropathologists (EBL and John Q. Trojanowski).[1]

Using consensus criteria,[2-5] ten pathologic features that reflect four proteinopathies (tau, amyloid-β, α-synuclein, TDP-43) and three cerebrovascular lesions (infarct, amyloid angiopathy, arteriolosclerosis) were obtained for each case. Specifically, amyloid-β positive plaques were assessed to determine Thal amyloid phase 0, 1/2, 3, and 4/5, corresponding to a score of 0, 1, 2, and 3. Tau-positive neurofibrillary tangles and neuritic plaques were assessed to determine Braak stage, 0, I/II, III/IV, and V/VI, corresponding to Braak score 0, 1, 2, and 3, and CERAD scores from 0 to 3, respectively.[3] These scores were used to determine the overall level of Alzheimer's disease neuropathologic change (ADNC) corresponding to not, low, intermediate, and high ADNC.[3] α-synuclein positive Lewy bodies and Lewy neurites were used to assign cases into no LBD versus brainstem, amygdala, limbic/transitional, or neocortical/diffuse LBD which was categorized into the absence or presence of LBD, where brainstem LBD was included in the absence group considering the lack of association of brainstem LBD with cognitive dysfunction.[2] The anatomical distribution of 1D3 positive TDP-43 inclusions determined the stages of limbic-predominant age-related TDP-43 encephalopathy neuropathologic change (LATE-NC; stage 0, no LATE; stage 1, amygdala; stage 2, hippocampus; stage 3, middle frontal cortex).[4] H&E and immunohistochemical staining of amyloid-β were assessed to determine three cerebrovascular lesions, absence or presence of large infarcts, absence or presence of moderate to high occipital cerebral amyloid angiopathy burden, and absence or presence of moderate to high occipital white matter arteriolosclerosis burden.[5] These cerebrovascular disease scores were then used to determine a low, intermediate, or high likelihood that cerebrovascular pathology contributed to cognitive impairment according to the Vascular Cognitive Impairment Neuropathology Guidelines (VCING) criteria for cerebrovascular disease.[5]

### Air pollution exposure assessment

We estimated annual concentrations of $PM_{2.5}$ at a 0.01° × 0.01° (approximately 1.1km x 1.1km) grid resolution across the US from 1998 through 2022 (V5.GL.04), which were recently updated with extended temporal coverage from previously described data sets (1998-2019) through 2022.[6] These data were derived from three sources, satellite-derived, chemical transport model simulation-based, and monitor based-information. Specifically, aerosol optic depth (AOD) was obtained from four satellites instruments, MODIS, VIIRS, MISR, and SeaWiFS, and their respective retrievals (Dark Target, Deep Blue, MAIAC), each daily AOD product was integrated with chemical transport model (GEOS-Chem) simulation based on biases and uncertainties of ground-based measurements of AOD from the global Aerosol Robotic Network (AERONET). These daily AOD products were then calibrated and combined to produce monthly and annual AOD at 0.01° × 0.01° (approximately 1.1km x 1.1km) grids using geographically weighted regression. These data sets were compared with ground-level $PM_{2.5}$ concentrations measured from monitoring stations (n=2312) across North America, with a cross-validated $R^2$ of 0.70 indicating good model performance.

We matched annual average estimates of $PM_{2.5}$ concentration data to the geocoded residential address of each case for the year of death. Specifically, we overlaid geocoded latitude and longitude coordinates of cases' residential locations on grid cells with raster values representing annual average $PM_{2.5}$ estimates and extracted raster values from grid cells at the geocoded point locations using a sample raster values tool in Quantum geographic information system (QGIS) software version 3.36.0. Finally, we obtained annual average $PM_{2.5}$ concentration in the year of death for each case as an exposure measure.

### Diagnostic statistical tests

We assessed the structural equation model fit using comparative fit index (CFI), standard root mean squared residual (SRMR), and Tucker-Lewis Index (TLI), with CFI greater than 0.95, SRMR less than 0.05, and TLI greater than 0.95 indicating a very good model fit.[7]

$PM_{2.5}$ measures are spatially distributed data such that data from locations close to each other may have more similar values than those further apart, which may consequently affect residuals of statistical models.[8] Thus, we further tested whether residuals from linear regressions were spatially autocorrelated or independent using Moran's Index (I) test.[9] The expected values of Moran's I range from -1 to +1. Negative values close to -1 indicate that similar residuals are from uniformly dispersed areas. Positive values close to +1 indicate that similar residuals are from areas close to each other. Values close to 0 indicate that residuals are from randomly distributed areas without spatial autocorrelation. A p value < 0.05 was set as statistical significance, with a null hypothesis of no residual spatial autocorrelation. In our adjusted linear regression model with one-year average exposure to $PM_{2.5}$ before death as the exposure variable and the Clinical Dementia Rating-Sum of Boxes (CDR-SB) score at last assessment as the outcome variable (Table 2), analysis of residual spatial autocorrelation did not reveal any spatial dependency for the linear regression model (expected I= -0.003; P=0.17). This indicates that the fitted model was well specified, suggesting that the observed association of $PM_{2.5}$ exposure with CDR-SB scores was independent from spatial patterns.

**Supplementary Figure 1. Flow chart of study participants**

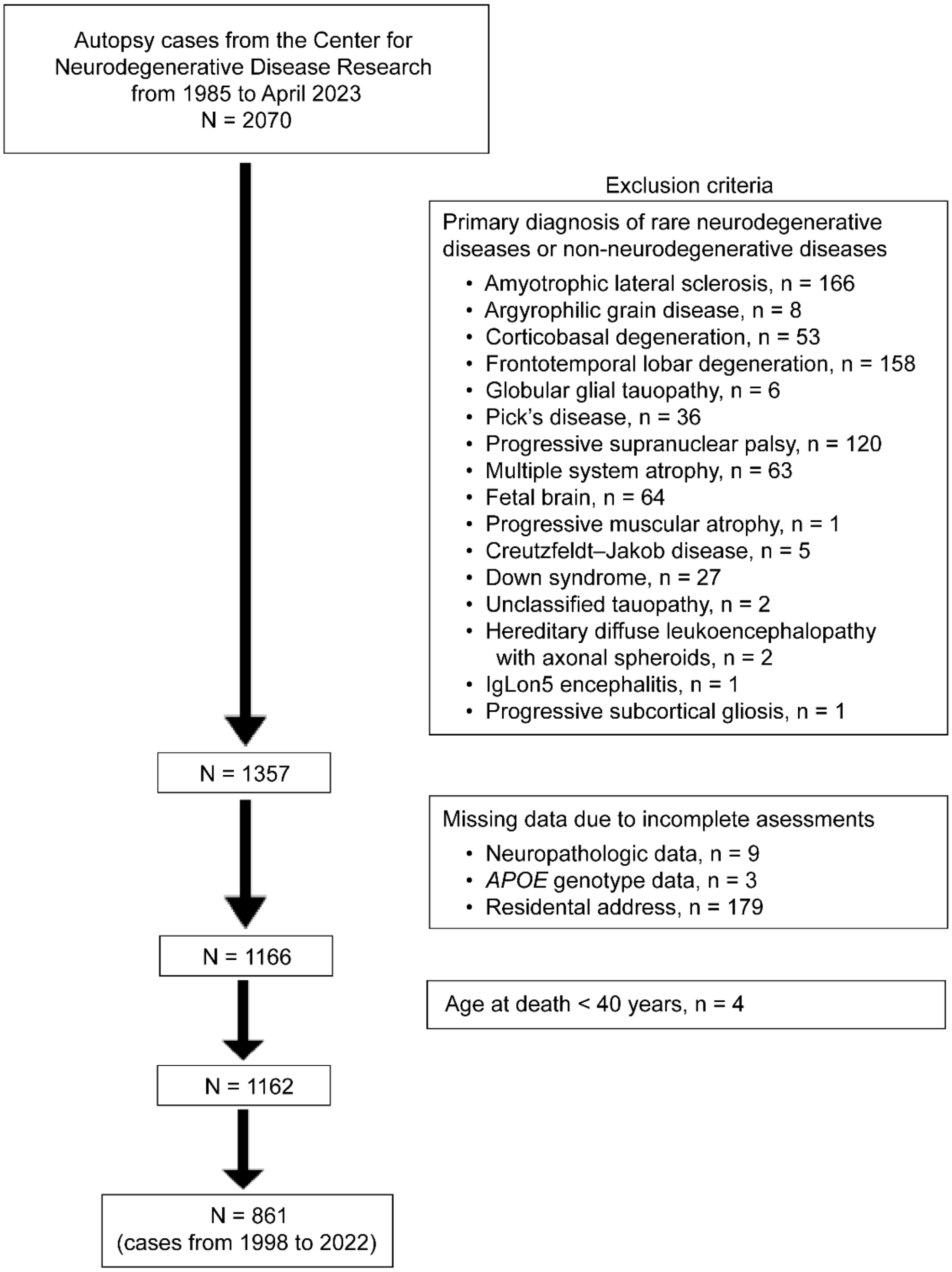

### Supplementary Figure 2. Spatial distribution of the study cohort with exposure to $PM_{2.5}$

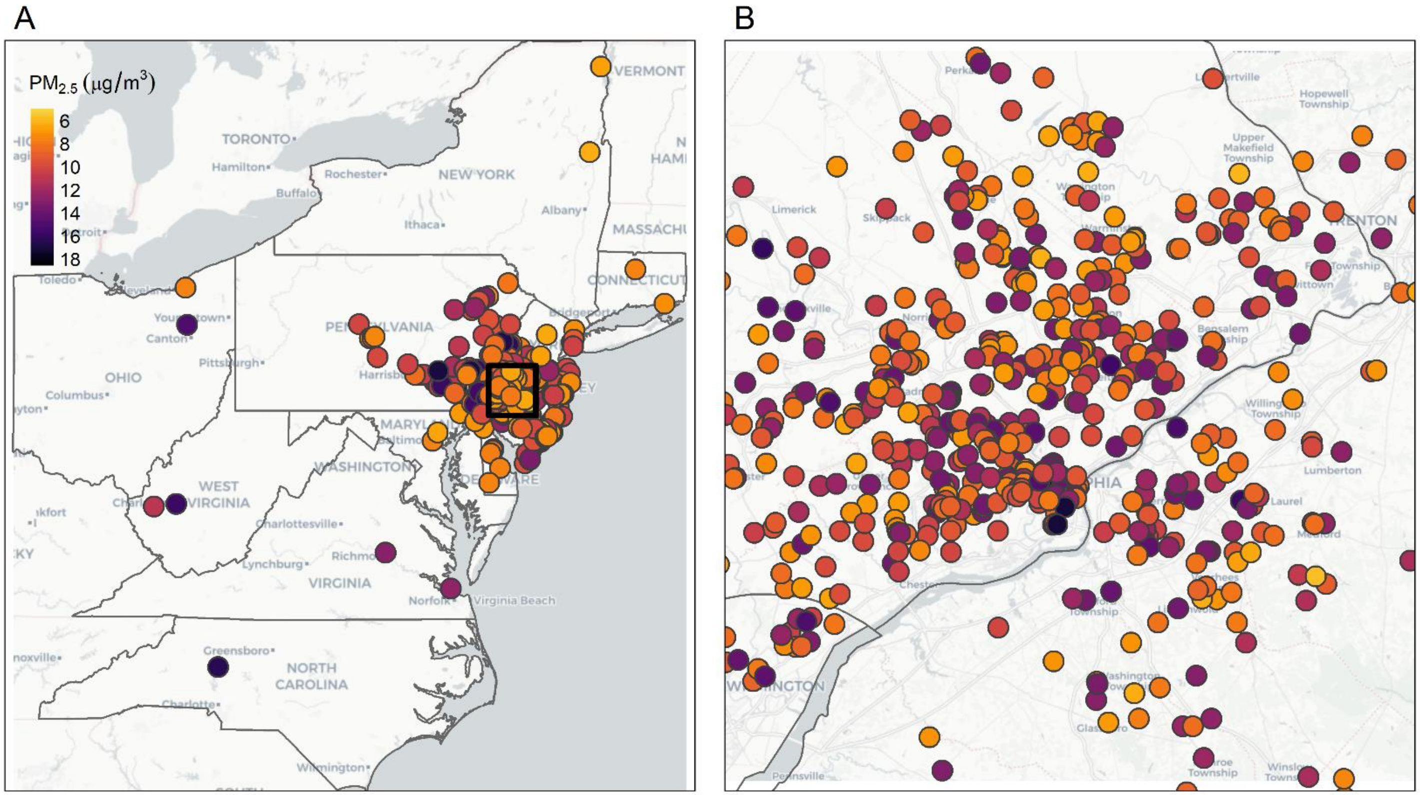

(A) Spatial distribution of study cases across ten states (n=858 of 861, 3 from California, Colorado, and Florida), color-coded by one-year average $PM_{2.5}$ concentration exposure before death. The rectangular box indicates Philadelphia city and its neighbouring regions. (B) Philadelphia city region corresponding to the residences of a large proportion of this autopsy cohort. Each dot provides data on a geocoded residential location and annual average $PM_{2.5}$ level (μg/$m^3$) for each case.

**Supplementary Figure 3. Cohort and national trends in $PM_{2.5}$ concentrations**

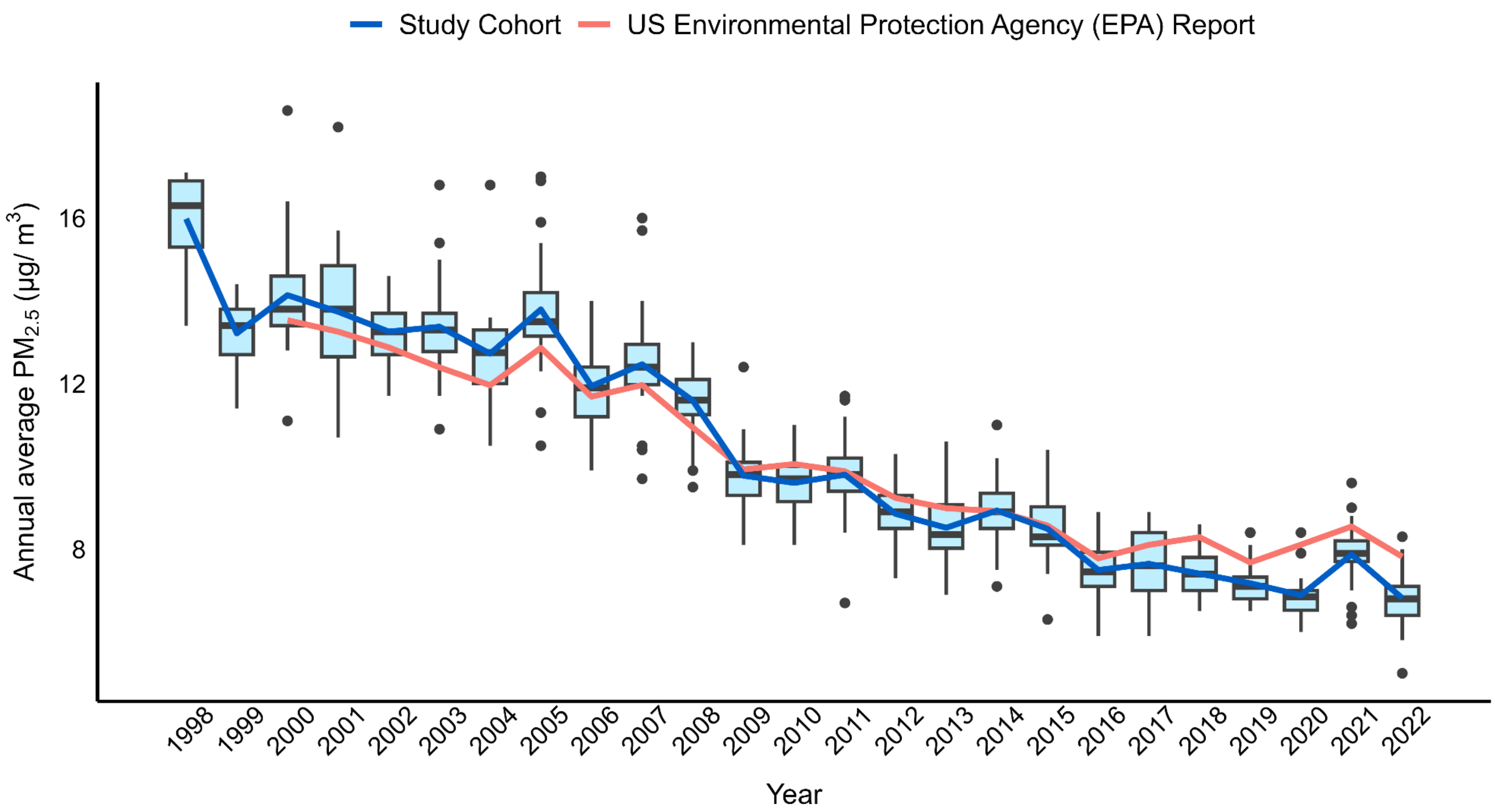

Annual average exposure to $PM_{2.5}$ concentrations for the year of death (one-year average exposure before death) for our study cohort (Study Cohort, blue line, box and whisker plots with points) from 1998 through 2022 and the annual average exposure to $PM_{2.5}$ concentrations for a nationwide network of 361 monitoring sites, reported by the US EPA from 2000 through 2022 (US EPA Report, orange line). In each box plot, the horizontal line within the box indicates the median value, the top and bottom edges of the box indicate the interquartile range, and the lower and upper whiskers indicate the lowest and highest values within 1.5 times the interquartile range of the bottom and top of the box, respectively. Points indicate outlier values outside the range. Annual mean $PM_{2.5}$ concentrations are provided for each group. $PM_{2.5}$ levels have decreased by 51.8% in our cohort (blue line) and 42.2% in the national cohort (orange line) from 2000 through 2022, showing a strong correlation of $PM_{2.5}$ trends (n=23, r = 0.97 and $p < 0.001$ by spearman's rank correlation).

**Supplementary Table 1. Different $PM_{2.5}$ exposure time windows and Clinical Dementia Rating–Sum of Boxes (CDR-SB).**

| | 1-year | | 3-year | | 5-year | |
|---|---|---|---|---|---|---|
| Exposure | Estimate (95% CI) | P Value | Estimate (95% CI) | P Value | Estimate (95% CI) | P Value |
| | No. of participants, N = 348† | | N = 347‡ | | N = 335§ | |
| $PM_{2.5}$ | 0.78 (0.52, 1.05) | <0.001 | 0.77 (0.5, 1.03) | <0.001 | 0.76 (0.49, 1.03) | <0.001 |
| Education | -0.31 (-0.53, -0.09) | 0.005 | -0.31 (-0.53, -0.09) | 0.006 | -0.3 (-0.53, -0.08) | 0.008 |

Abbreviation: CI, confidence interval.

Associations were evaluated by three linear regression models in which the outcome variable was the Clinical Dementia Rating-Sum of Boxes (CDR-SB) score at last assessment where higher scores indicating greater cognitive and functional impairment, and exposure variables were years of education and either 1-year, 3-year, or 5-year $PM_{2.5}$ exposure before death. All models were controlled for sex, age at last CDR-SB assessment, and race.

1-year model is the main model presented in Table 2.

†N=Number of autopsy cases with CDR-SB data from 1998 through 2022. ‡N= Number of autopsy cases with CDR-SB data from 2000 through 2022. §N=Number of autopsy cases with CDR-SB data from 2002 through 2022.

Estimated effects and 95% confidence intervals are shown as adjusted CDR-SB score at last assessment prior to death per 1 $\mu g/m^3$ increase in 1-year, 3-year, and 5-year average $PM_{2.5}$ exposure before death and one-year increase in educational attainment, with positive values indicating greater cognitive and functional impairment.

P values less than 0.05 were considered statistically significant.

**Supplementary Table 2. Different $PM_{2.5}$ exposure time windows and longitudinal change in Clinical Dementia Rating–Sum of Boxes (CDR-SB).**

| | 1-year | | 3-year | | 5-year | |
|---|---|---|---|---|---|---|
| Interaction term | Estimate (95% CI) | P Value | Estimate (95% CI) | P Value | Estimate (95% CI) | P Value |
| | No. of participants, N=263[†] | | N=263[‡] | | N=257[§] | |
| $PM_{2.5}$ and Interval years between CDR-SB assessment and death | 0.13 (0.09, 0.16) | <0.001 | 0.15 (0.12, 0.19) | <0.001 | 0.15 (0.12, 0.18) | <0.001 |

Abbreviation: CI, confidence interval.

Associations were evaluated with three linear mixed-effects models in which the outcome variable was annual change in the Clinical Dementia Rating-Sum of Boxes (CDR-SB) scores obtained from multiple assessments where higher values indicating faster cognitive and functional impairment, and the exposure variable was the interaction term between 1-year, 3-year, or 5-year average $PM_{2.5}$ exposure before death and interval years between last CDR-SB assessment and death. All models were controlled for interval years between last CDR-SB assessment and death, sex, age at last CDR-SB assessment, race, and years of education.

1-year model is the main model presented in Table 3.

†N=Number of autopsy cases with multiple CDR-SB data from 1998 through 2022. ‡N= Number of autopsy cases with multiple CDR-SB data from 2000 through 2022. §N=Number of autopsy cases with multiple CDR-SB data from 2002 through 2022.

Estimated effects and 95% confidence interval are shown as adjusted annual change in CDR-SB score at last assessment prior to death per 1 $\mu g/m^3$ increase in 1-year, 3-year, and 5-year average $PM_{2.5}$ exposure before death, with positive values indicating faster cognitive and functional impairment.

P values less than 0.05 were considered statistically significant.

### Supplementary Table 3. Different $PM_{2.5}$ exposure time windows and dementia-related neuropathologic change

| | 1-year | | 3-year | | 5-year | |
|---|---|---|---|---|---|---|
| Outcome | Odds Ratio (95% CI) | P Value | Odds Ratio (95% CI) | P Value | Odds Ratio (95% CI) | P Value |
| | No. of participants, N = 861† | | N = 823‡ | | N = 778§ | |
| Thal amyloid phase | 1.07 (1.01, 1.13) | 0.02 | 1.10 (1.04, 1.16) | 0.002 | 1.11 (1.04, 1.18) | 0.001 |
| Braak stage | 1.07 (1.02, 1.13) | 0.007 | 1.10 (1.04, 1.17) | 0.001 | 1.12 (1.05, 1.18) | <0.001 |
| CERAD score | 1.11 (1.05, 1.17) | <0.001 | 1.13 (1.07, 1.19) | <0.001 | 1.15 (1.08, 1.22) | <0.001 |
| ADNC level | 1.06 (1.01, 1.12) | 0.02 | 1.10 (1.04, 1.16) | 0.001 | 1.11 (1.05, 1.18) | <0.001 |
| LBD Stage | 1.01 (0.96, 1.06) | 0.82 | 1.01 (0.96, 1.07) | 0.71 | 1.01 (0.95, 1.07) | 0.85 |
| LATE-NC | 1.02 (0.96, 1.08) | 0.52 | 1.02 (0.96, 1.09) | 0.50 | 1.04 (0.97, 1.11) | 0.23 |
| Large infarct | 1.17 (1.05, 1.30) | 0.005 | 1.18 (1.04, 1.33) | 0.009 | 1.16 (1.02, 1.33) | 0.03 |
| Cerebral amyloid angiopathy | 0.98 (0.92, 1.04) | 0.52 | 0.97 (0.91, 1.04) | 0.42 | 0.93 (0.87, 1.00) | 0.04 |
| Arteriolosclerosis | 0.99 (0.92, 1.06) | 0.78 | 1.00 (0.93, 1.08) | 0.94 | 0.96 (0.88, 1.04) | 0.29 |
| VCING level | 1.05 (0.97, 1.14) | 0.25 | 1.06 (0.97, 1.16) | 0.20 | 1.02 (0.92, 1.12) | 0.75 |

Abbreviations: CERAD, Consortium to Establish a Registry for Alzheimer’s Disease; ADNC, Alzheimer’s disease neuropathologic change; LBD, Lewy body disease; LATE, limbic-predominant age-related TDP-43 encephalopathy; VCING, vascular cognitive impairment neuropathology guidelines; CI, confidence interval.

Associations were calculated as odds ratios and 95% confidence intervals for neuropathologic outcomes corresponding to every 1 $\mu g/m^3$ increase in 1-year, 3-year, and 5-year average $PM_{2.5}$ exposure before death.

1-year model is the main model presented in Figure 1.

†N=Number of autopsy cases from 1998 through 2022. ‡N= Number of autopsy cases from 2000 through 2022. §N=Number of autopsy cases from 2002 through 2022.

Odds ratios and 95% confidence intervals are reported from a series of ordinal logistic regression models with 1-year, 3-year, or 5-year average $PM_{2.5}$ exposure before death as the exposure variable and ordinal Thal amyloid phase, ordinal Braak stage, ordinal CERAD score, ordinal ADNC level, ordinal LATE-NC stage, and ordinal VCING score as outcome variables, and binary logistic regression models with 1-year, 3-year, or 5-year average $PM_{2.5}$ exposure before death as the exposure variable and dichotomously treated LBD stage, presence of large infarcts, presence of occipital cerebral amyloid angiopathy, and presence of arteriolosclerosis as outcome variables. Odds ratios and 95% confidence intervals greater than 1 indicate worse neuropathologic outcomes. All models were controlled for sex, age at death, and *APOE* ε4 status.

P values less than 0.05 were considered statistically significant.

**Supplementary Table 4. Mediation Analysis of Dementia-Related Neuropathologic Change on the Association between Different PM2.5 Exposure Time Windows and CDR-SB (Sensitivity Analysis)**

| | 1-year | | 3-year | | 5-year | |
|---|---|---|---|---|---|---|
| Effect | Estimate | 95% CI | Estimate | 95% CI | Estimate | 95% CI |
| | No. of participants, N = 348† | | N = 347‡ | | N = 335§ | |
| Indirect, ADNC level | 0.36 | 0.13, 0.65 | 0.34 | 0.12, 0.64 | 0.32 | 0.10, 0.61 |
| Indirect, Large infarct | 0.11 | -0.11, 0.87 | 0.18 | -0.03, 1.29 | 0.21 | -0.06, 1.06 |
| Direct | 0.10 | -0.48, 0.72 | 0.04 | -0.66, 0.67 | 0.04 | -0.73, 0.71 |
| Total | 0.57 | 0.35, 0.81 | 0.57 | 0.33, 0.80 | 0.57 | 0.34, 0.81 |

Abbreviations: ADNC, Alzheimer's disease neuropathologic change; CI, confidence interval.

Associations were evaluated using structural equation modeling for relationships between different $PM_{2.5}$ exposure time frames, ADNC or the presence of large infarcts, and CDR-SB with 1-year, 3-year, or 5-year average $PM_{2.5}$ exposure before death as the exposure variable, ordinal ADNC level and presence of infarcts as parallel mediators, and CDR-SB at last assessment as the outcome variable.

1-year model is the main model presented in Figure 2.

†N=Number of autopsy cases with CDR-SB data from 1998 through 2022. ‡N= Number of autopsy cases with CDR-SB data from 2000 through 2022. §N=Number of autopsy cases with CDR-SB data from 2002 through 2022.

Indirect effects are the effect of $PM_{2.5}$ exposure on CDR-SB through either ADNC or the presence of large infarcts, and the direct effect is the effect of $PM_{2.5}$ exposure on CDR-SB that is independent from mediators such as ADNC or the presence of large infarcts. All models were controlled for sex, age at death, interval years between last CDR-SB assessment and death, race, years of education, and *APOE* ε4 status.

Estimates of indirect, direct, and total effects with 95% confidence intervals from a mediation analysis with structural equation modeling using a bias-corrected and accelerated bootstrap confidence interval method with 2000 simulations. Positive values indicate greater cognitive and functional impairment.

All models exhibited acceptable fit indices (1-year model, comparative fit index [CFI]=0.895, standardized root mean squared residual [SRMR]=0.088, Tucker-Lewis index [TLI]=0.965; 3-year model, CFI=0.909, SRMR=0.091, TLI=0.970; 5-year model, CFI=0.907, SRMR=0.082, TLI=0.969).
